# Accelerated DMN-Targeted cTBS Improves Processing Speed Deficits in Schizophrenia

**DOI:** 10.64898/2026.02.11.26346103

**Authors:** Jillian G. Connolly, Sophia H. Blyth, Gulcan Yildiz, Baxter P. Rogers, Simon Vandekar, Mark A. Halko, Roscoe O. Brady, Heather Burrell Ward

## Abstract

**Objective:** Cognitive deficits are a leading cause of disability in schizophrenia and are linked to poor functional outcomes. There are no first line treatments for these deficits, and their neural basis is poorly understood. While schizophrenia is associated with widespread cognitive deficits, information processing speed is most profoundly impaired. Processing speed deficits have been associated with hyperconnectivity in the Default Mode Network (DMN). We therefore tested if modulating DMN connectivity with single or multiple sessions of transcranial magnetic stimulation (TMS) applied to an individualized DMN target would affect processing speed.

**Methods:** In the first study, 10 individuals with schizophrenia received single TMS sessions and underwent resting-state neuroimaging and processing speed assessment (Brief Assessment of Cognition in Schizophrenia digit symbol coding) acutely before and after each session. These sessions included excitatory (intermittent theta burst stimulation, iTBS); inhibitory (continuous theta burst stimulation, cTBS); and sham stimulation sessions. In the second study, 29 individuals (17 schizophrenia, 12 non-psychosis controls) received 5 accelerated sessions of cTBS with resting-state neuroimaging and processing speed assessment before and after the course of TMS sessions.

**Results:** In the accelerated, multi-session DMN-targeted TMS trial, cTBS improved processing speed in the schizophrenia group (p=0.0124). In individuals with schizophrenia, reduction in DMN connectivity was linked to improvement in processing speed (p=0.021). These changes were dependent on age, where younger participants experienced greater processing speed improvements than older participants (p=0.006).

**Conclusions:** In sum, personalized network targeted TMS is a novel method for reducing cognitive impairment associated with schizophrenia.

## Introduction

Cognitive deficits are a leading cause of disability in schizophrenia, contributing to poor occupational, educational, and social outcomes. People with schizophrenia are 51% less likely to enter higher education compared to the general population, and only 42.9% of adults with schizophrenia hold a full-time job (1,2). Although progress has been made in advancing the treatment of positive and negative symptoms of schizophrenia, there are currently no approved treatments for cognitive impairment in schizophrenia (3).

Processing speed is fundamental to the cognitive impairments observed in schizophrenia (4). The hierarchical model of cognition views processing speed as an underlying cognitive process that supports higher-level cognitive processes such as working memory and executive function (5).

Previous research found that differences in attention, verbal memory, visual memory, working memory, and executive functioning between schizophrenia and a control sample were lessened after controlling for processing speed (6). This finding suggests that underlying processing speed deficits may evoke downstream cognitive impairments in schizophrenia, making processing speed an optimal intervention target due to its foundational role in other cognitive domains.

The neural circuitry associated with processing speed dysfunction in schizophrenia is not well understood. Previous research suggests that Default Mode Network (DMN) hyperconnectivity is associated with an inability to properly allocate attentional resources in schizophrenia, leading to general cognitive impairments (7). The dysconnectivity hypothesis of schizophrenia links general cognitive impairments to abnormal neuronal activity in networks such as the DMN (8,9). In an early psychosis sample, evidence indicates that an inability to suppress the DMN during cognitive tasks is related to slowed processing speed (10). Beyond early psychosis, increased DMN activity in individuals with schizophrenia, schizoaffective disorder, and their first-degree relatives has been associated with decreased working memory performance (11) which is known to be directly affected by processing speed (12). It is well established that DMN hyperactivity is associated with cognitive impairment in schizophrenia. Taken together, current research suggests that the DMN may be an ideal target for potential treatments of processing speed impairment in schizophrenia.

Neuromodulation interventions can be used to noninvasively and selectively target the DMN. Transcranial Magnetic Simulation (TMS) is a form of non-invasive brain stimulation that uses a magnetic coil to stimulate or suppress brain activity in a targeted region (13). Given the well-established links between DMN hyperconnectivity and cognitive deficits in schizophrenia, we used a series of TMS experiments applied to a personalized DMN target to test if using TMS to reduce DMN connectivity would improve processing speed. We first applied single sessions of DMN-targeted TMS using a within-subjects, randomized crossover design of three different TMS protocols: intermittent theta-burst stimulation (iTBS), continuous theta-burst stimulation (cTBS), and sham. However, single sessions of TMS did not affect processing speed. Therefore, we scaled up the intervention to test if multiple sessions of DMN-targeted cTBS delivered in an accelerated protocol would improve processing speed. We hypothesized that DMN-targeted cTBS would improve processing speed in the schizophrenia group and that reduced DMN connectivity would be linked to processing speed improvements.

However, age is also known to play a crucial role in cognitive functioning in schizophrenia. Longitudinal research revels expedited cognitive decline over the lifetime in schizophrenia compared to general psychosis or control populations (14)(15). Therefore, younger individuals may be more receptive to cognitive interventions than older populations with schizophrenia. As an exploratory hypothesis, we predicted that younger participants would experience greater cognitive improvements following cTBS than older participants. In the control group, we hypothesized that DMN-targeted cTBS would not improve processing speed and that reduced DMN connectivity would be linked to reductions processing speed.

## Methods

### Single-Session DMN-Targeted TMS in Schizophrenia

#### Participants

Fifteen individuals with schizophrenia or schizoaffective disorder aged 18-65 were enrolled in this randomized, sham-controlled, crossover TMS-fMRI study (Figure 1A, Table 1, Supplemental Figure 1, NCT07190352). Participants provided written informed consent in accordance with the Beth Israel Deaconess Medical Center Institutional Review Board. Details in supplement.

**Figure 1.**
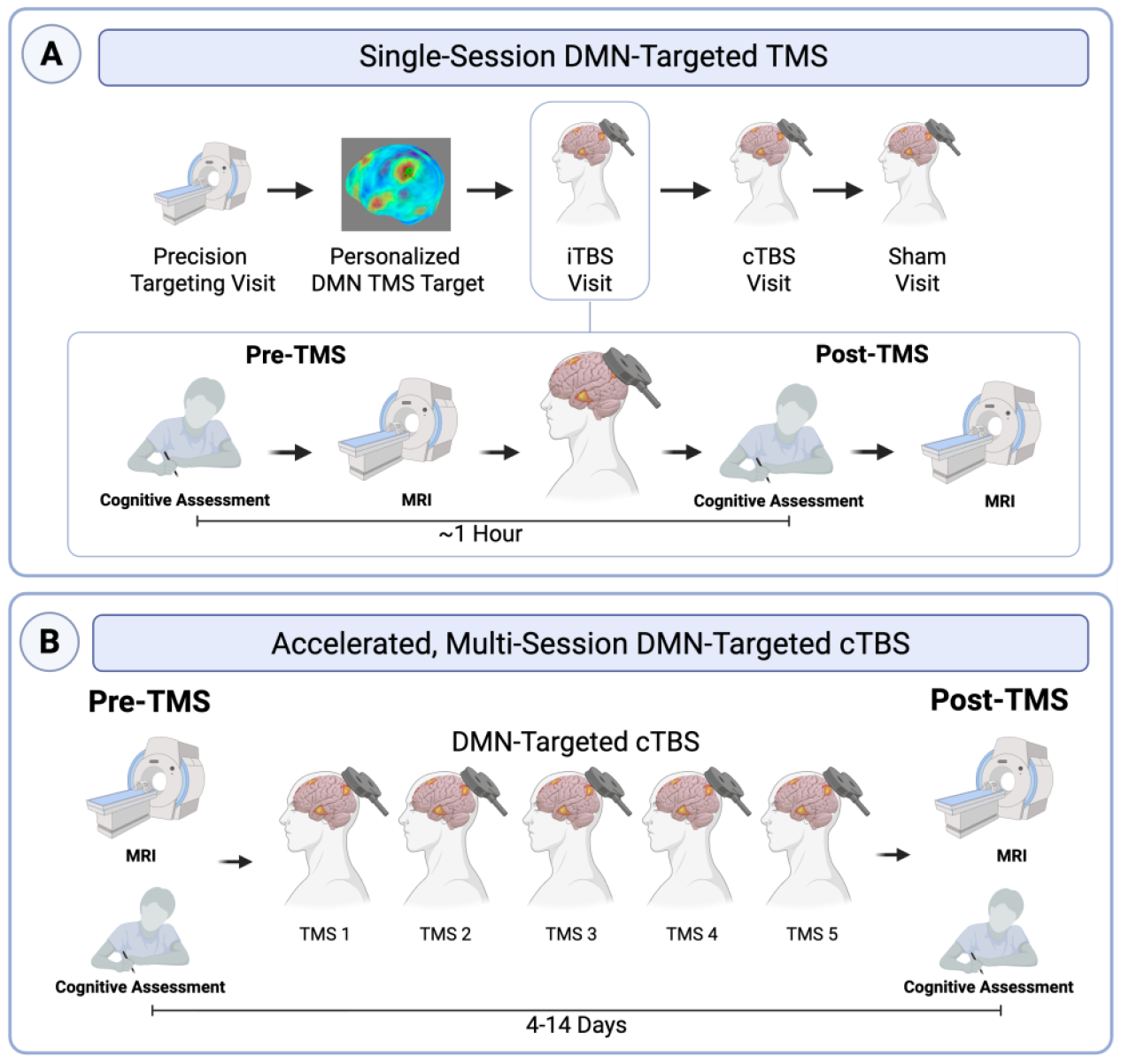
Study design comparison for Single-Session DMN-Targeted TMS compared to Accelerated, Multi-Session DMN-Targeted cTBS. In the Single-Session DMN-targeted TMS study, 10 individuals with schizophrenia received single TMS sessions (intermittent theta burst stimulation, iTBS; continuous theta burst stimulation, cTBS; sham) and underwent resting-state neuroimaging and processing speed assessment (Brief Assessment of Cognition in Schizophrenia digit symbol coding) before and after each session. Approximately one hour passed between pre-TMS and post-TMS cognitive testing. In the Accelerated, Multi-Session DMN-targeted cTBS study, 29 individuals (17 schizophrenia, 12 non-psychosis controls) received 5 accelerated sessions of cTBS with resting-state neuroimaging and processing speed assessment before and after TMS. Approximately 7 days passed between pre-TMS and post-TMS testing (median 7 days, mean 7.79 days (SD 2.57), range 4-14 days).

**Figure 2.**
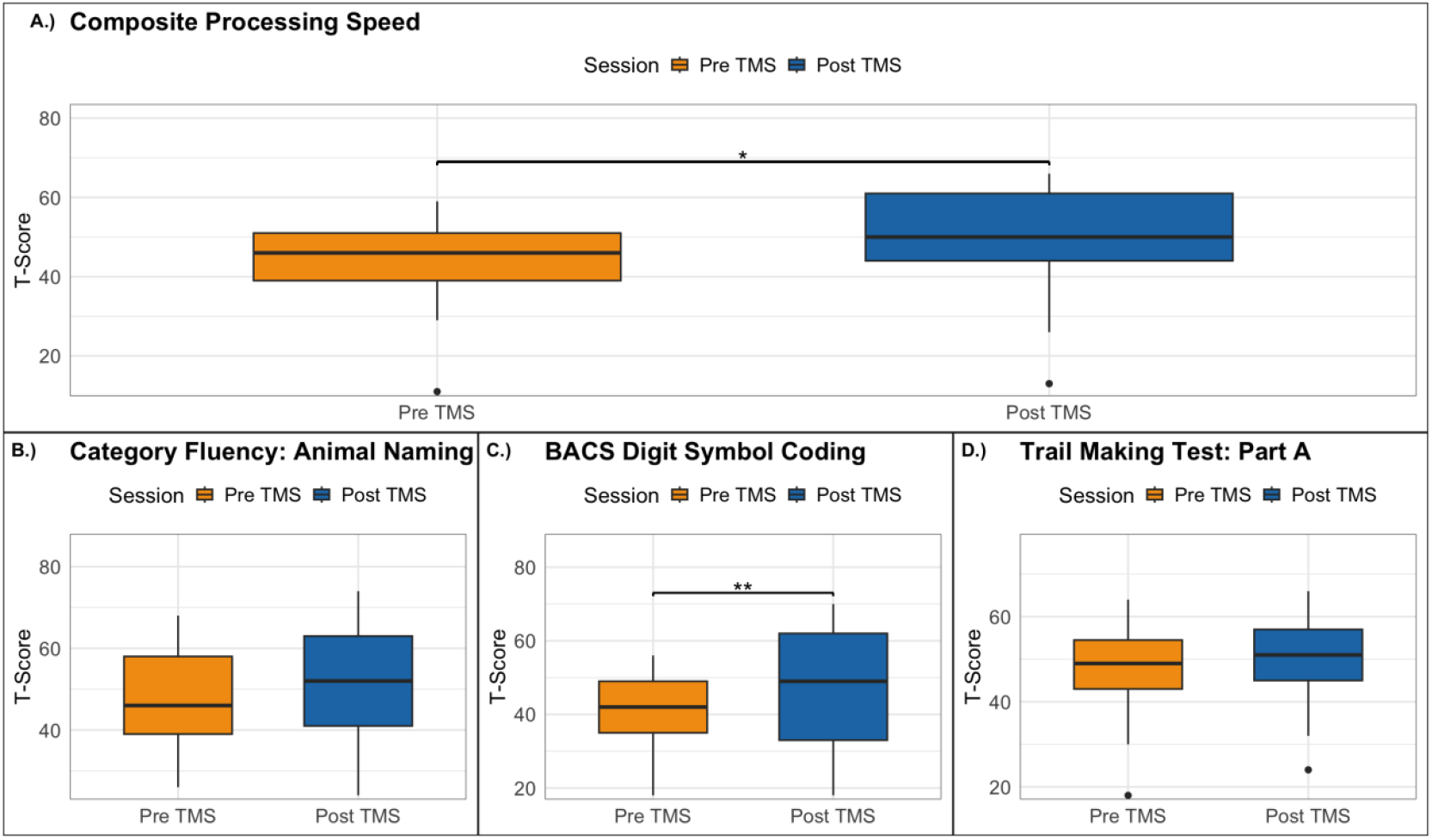
Accelerated, Multi Session cTBS improves overall processing speed in schizophrenia. Seventeen individuals with schizophrenia received five accelerated sessions of DMN-targeted cTBS with pre-post assessment of processing speed. Processing speed was assessed using three individual tests: 1) Category Fluency: Animal Naming; 2) BACS Digit Symbol Coding and 3) Trail Making Test (TMT): Part A, which were compiled to create a composite processing speed t-score corrected for age and gender. Composite processing speed score (p=0.0124, Figure 1A) and Digit Symbol Coding score (p=0.0083, Figure 1C) significantly improved in the schizophrenia group. Animal Fluency (p=0.274, Figure 1B) and TMT (p=0.382, Figure 1D) revealed non-significant numerical increases in median.

**Figure 3.**
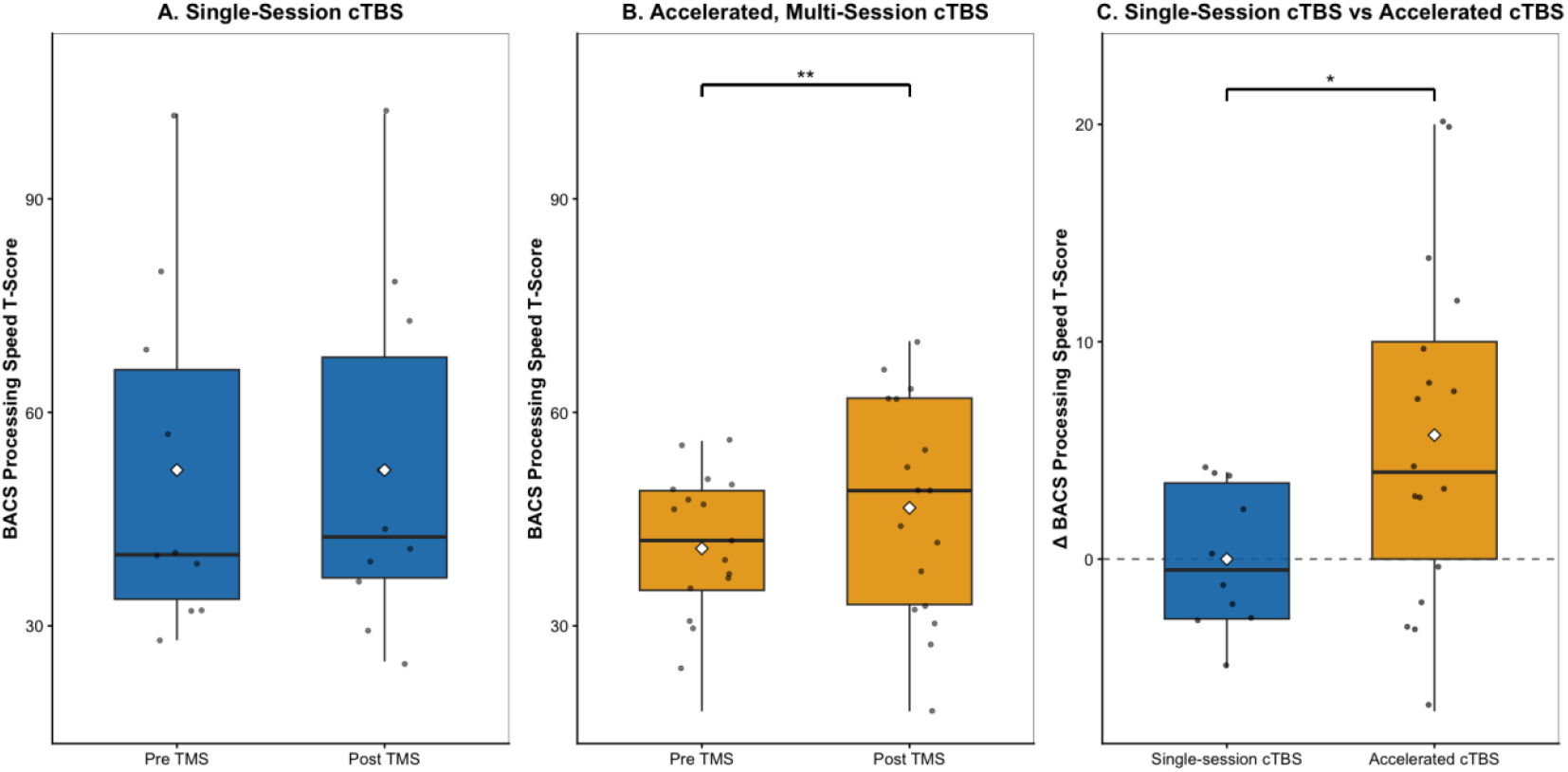
Accelerated, multi-session cTBS improves processing speed in schizophrenia significantly more than single-session cTBS. Ten individuals with schizophrenia received single sessions of DMN-targeted cTBS with pre-post processing speed assessment. Single sessions of cTBS did not affect processing speed in schizophrenia (p=1.00, Figure 2A). Seventeen individuals with schizophrenia received five accelerated sessions of DMN-targeted cTBS with pre-post assessment of processing speed. Accelerated multi-session DMN-targeted cTBS improved processing speed performance (p=0.0124, Figure 2B). When the effects of single- and multiple sessions of DMN-targeted cTBS were directly compared multi-session DMN-targeted cTBS showed significantly higher processing speed change scores than with single session cTBS (p=0.0148, Figure 2C).

#### Cognitive Assessments

Prior to receiving TMS, the Brief Assessment of Cognition in Schizophrenia Digit Symbol Coding (BACS processing speed) task was administered to all participants. The BACS processing speed task is a sub-test of the processing speed domain of the Measurement and Treatment Research to Improve Cognition in Schizophrenia (MATRICS) (16). Raw scores were transformed to a corrected t-score to account for age and gender (17). After TMS administration (i.e., approximately two hours after the first test), participants repeated the BACS Digit Symbol Coding task. Details in supplement.

#### Individualized DMN Targeting

For each participant, an individualized DMN target was identified in the left posterior inferior parietal lobule (IPL). This DMN region was selected given evidence demonstrating TMS applied to a personalized left IPL DMN target modulates DMN connectivity (18). To generate an individualized DMN map for TMS targeting, a standard DMN template (Yeo et al., 2011) was warped into native space and applied to the participant’s pre-TMS scan to obtain a maximally correlated DMN IPL target. In each participant, the resultant connectivity maps yielded a correlation cluster in the left posterior IPL. A target was placed in the averaged center of the left posterior IPL correlation cluster (formed from the overlay of the left posterior IPL clusters derived from connectivity maps) on the cortical surface using Brainsight neuronavigation software (Supplemental Figure 3). This individualized TMS target in the left IPL region of the DMN was used as the TMS target for all sessions in both Single-Session DMN-Targeted TMS and Accelerated, Multi-Session DMN-Targeted cTBS.

#### TMS Protocol

Individuals received single sessions of theta-burst stimulation applied to an individualized DMN target with neuroimaging collected immediately before and after each session. Individuals received one session of iTBS (600 pulses, 100% Active Motor Threshold, AMT), cTBS (600 pulses, 80% AMT as per (19)), and sham (coil flipped 180 degrees, 100% AMT iTBS protocol, 600 pulses) on three separate days, separated by at least 2 days to avoid carryover effect (median 6.5 days, mean 13.8 days (SD 22.2), range 2-96 days). Order was randomized, and participants were blinded to stimulation type. Details in supplement.

#### MRI Acquisition & Processing

Imaging data were collected on a Siemens 3.0-T MRI system (Munich, Germany). Participants each completed 7 MRI scans, including at baseline and immediately before and after each TMS session (Figure 1B). A total of 270 min of imaging data was collected per participant, including 140 min of resting-state imaging. Briefly, 1-mm^3^ T1-weighted anatomical scans and multiple 10-minute functional runs were acquired. All resting-state scans went through a quality assurance procedure that included calculating framewise displacement (FD) and temporal signal to noise ratio (tSNR). Scans with mean FD>0.5 or tSNR<5th percentile of the sample distribution were excluded. Details in supplement.

### Accelerated, Multi-Session DMN-Targeted cTBS in Schizophrenia

#### Participants

Eighteen individuals with schizophrenia or schizoaffective disorder and thirteen non-psychosis control subjects aged 18-65 were enrolled in this TMS-fMRI study (Figure 1B, Table 1, Supplemental Figure 2, NCT07155096). Participants provided written informed consent in accordance with the Vanderbilt University Medical Center IRB. Details in supplement.

#### Cognitive Assessment

Participants completed three tasks comprising the MATRICS Processing Speed Domain including BACS Digit Symbol Coding (BACS processing speed), Category Fluency: Animal Naming, and Trail Making Test: Part A. The MATRICS online scoring program provided T-scores for each sub-test as well as a composite Processing Speed T-score derived from a participant’s overall performance on the three tasks. Participants completed the tasks during the pre-TMS and post-TMS MRI visits, with at least four days in between tests (median 7 days, mean 7.79 days (SD 2.57), range 4-14 days). Details in supplement.

#### Accelerated, Multi-Session TMS Protocol

Individuals received 5 sessions of cTBS (600 pulses, 100% AMT, Figure 1B) applied to an individualized DMN target (see *Individualized DMN Target* above) in a one-day accelerated protocol (30-minute inter-session interval) with pre-/post-TMS neuroimaging. Because we intended to *reduce* DMN connectivity, cTBS was selected as the stimulation type for the following reasons: 1) cTBS has been associated with inhibitory effects (20); 2) iTBS applied to a DMN target increases DMN connectivity(21); and 3) we observed a greater numerical reduction in DMN connectivity following cTBS in our single session study compared to iTBS. TMS was applied using a MagPro X100 stimulator and an active figure-of-8 coil (Cool B65, MagVenture, Denmark) as above.

#### MRI Acquisition & Processing

Imaging data were collected on 3.0-T Philips Intera Achieva MRI scanner (Philips Healthcare, Andover, MA) before and after the TMS intervention. As the durability of 5 TMS sessions is unknown, post-TMS scans were scheduled 3-7 days after TMS per protocol. Briefly, 1-mm^3^ T1-weighted anatomical scans and multiple 10-minute functional runs were acquired. All resting-state scans went through a quality assurance procedure that included calculating FD and tSNR. Scans with mean FD > 0.6 or tSNR lower than the 5th percentile of the sample distribution were excluded from analysis. Details in supplement.

### TMS Neuroimaging Analysis - Calculation of TMS Target DMN Connectivity Values

For both TMS studies, we calculated average connectivity to the DMN from the TMS target (left parietal DMN) by extracting time courses from eight standard DMN nodes (22) and averaging connectivity between the TMS target and the eight other DMN nodes for each participant. We calculated individual values of DMN functional connectivity by placing 6mm spheres at coordinates corresponding to standard DMN nodes ((22) for coordinates). The BOLD signal time courses from the TMS target (left lateral parietal DMN) and the other DMN regions of interest (ROIs) were correlated with each other and z-transformed to generate ROI-to-ROI connectivity values. The mean DMN connectivity from the TMS target was generated for each participant by averaging connectivity values of these eight network edges. Details in supplement.

### Statistical Analysis

For the Single-Session DMN-targeted TMS study, paired t-tests were used to compare BACS processing speed t-scores between pre-TMS and post-TMS timepoints for the iTBS, cTBS, and sham conditions. A linear mixed effects model was used to compare the BACS processing speed change scores between cTBS, iTBS, and sham.

For the Accelerated, Multi-Session DMN-targeted cTBS study, processing speed was analyzed using paired t-tests between the pre and post t-scores for the BACS processing speed task, Animal Naming task, Trail Making task, and the overall composite processing speed score. A linear mixed effects model was also used to examine the interaction, if any, between group (control and schizophrenia) and time (pre-TMS and post-TMS) in relation to composite processing speed t-score change using the following structure: processing speed ∼ time + group + (1 | subject). BACS processing speed t-score change was compared for each single session intervention (cTBS, iTBS, and sham) against the accelerated, multi-session cTBS using three independent two-sample t-tests. To test for an age effect on processing speed change, the following linear model was used: processing speed change ∼ age + group. We used a linear regression model to test if DMN connectivity change, group, and their interaction were associated with processing speed change while controlling for age.

## Results

### Single Session TMS Does Not Significantly Affect Processing Speed in Schizophrenia

Ten individuals with schizophrenia completed the single session TMS randomized, controlled crossover study and were included in analysis. No serious adverse events were reported. Processing speed did not change after the administration of iTBS (t(9) = -0.6784, p = 0.514), cTBS (t(9) = 0, p=1.00), or sham (t(9) = -0.1098, p= 0.915), as measured by the BACS processing speed t-score. A linear mixed effects model revealed no significant effects of TMS type (iTBS: t(26.82) = 0.497, p= 0.62; cTBS: t(18) = -0.362, p = 0.722); sham: t(18) = -0.241, p = 0.812) on BACS processing speed.

There was no effect of TMS type on connectivity from the TMS target to DMN (iTBS: t(21)=-0.072, p=00.943; cTBS: t(21)= -0.410, p=0.686; sham: t(21)=0.800, p=0.432).

### Accelerated, Multi-Session cTBS Significantly Improves Processing Speed in Schizophrenia

Seventeen individuals with schizophrenia and twelve non-psychosis control participants completed this accelerated, multi-session DMN-targeted cTBS study and were included in analysis. No serious adverse events were reported.

Accelerated, multi-session DMN-targeted cTBS significantly improved composite processing speed in the schizophrenia group (t(16)=2.8176, p=0.0124, d=0.365). BACS processing speed t-score also significantly increased in the schizophrenia group (t(16)=3.0098, p=0.0083, d=0.363). However, performance on the other two subtests of MATRICS processing speed, Animal Fluency (t(16)= 1.132, p=0.274, d=0.215) and Trail Making Test, Part A (t(15)=0.902, p=0.382, d=0.1978), only revealed a non-significant numerical increase in mean score in the schizophrenia group.

In the control group, multi-session, accelerated cTBS improved overall processing speed (t(11)= 3.653, p=0.0038, d=0.745). BACS processing speed t-scores (t(11)= 2.699, p = 0.0207, d = 0.517) and Trail Making t-scores (t(11)=4.013, p=0.002, d=0.643) also significantly improved. However, Animal Fluency t-score did not significantly change after cTBS (t(11)= -0.658, p=0.524, d=-0.098).

We fit a linear mixed-effects model with composite processing speed t-score as the outcome to test for an interaction between group and time. We did not observe a group by time interaction (t(29) = 0.495, p=0.624). However, when we removed the interaction from the model, we found a significant main effect of time (t(29)=4.136, p = 0.0003) on processing speed score, where participants improved following TMS treatment. We also observed a main effect of group (t(29)= -3.166, p=0.0036) on processing speed, where participants with schizophrenia demonstrated significantly lower processing speed scores compared to the non-psychosis controls.

### Accelerated, Multi-Session cTBS Improves Processing Speed Significantly more than Single-Session cTBS in Schizophrenia

To directly compare the effects of single and multiple sessions of TMS on processing speed, we included the Single-Session DMN-targeted TMS and Accelerated, Multi-Session DMN-targeted cTBS groups in the same model and compared BACS processing speed t-score change between the two interventions. Accelerated, multi-session cTBS improved BACS processing speed t-scores significantly more than single session cTBS in the schizophrenia group (t(23.44)= 2.631, p=0.0148, d= 0.869). Accelerated cTBS did not change BACS processing speed t-scores significantly more than single session iTBS (t(23.85)=1.734, p=0.0951, d=0.635) or sham (t(13.959)=1.292, p=0.2173, d=0.569).

### Processing Speed Change After Accelerated, Multi-Session cTBS is Dependent on Age and Group

In a linear regression model examining processing speed change based on age, group, and the age*group interaction, there was a significant interaction between age and group (Estimate=-0.57, t(25)=-3.003 SE=0.19, p=0.0056) in relation to processing speed change. In the schizophrenia group, processing speed performance decreased with higher age, while in the control group age was not related to processing speed change (Figure 4). After removing the interaction term from the model, we did not observe a significant main effect of group (t(26) =0.065, p = 0.949) and age (t(26) = -1.632, p = 0.114) on processing speed.

**Figure 4.**
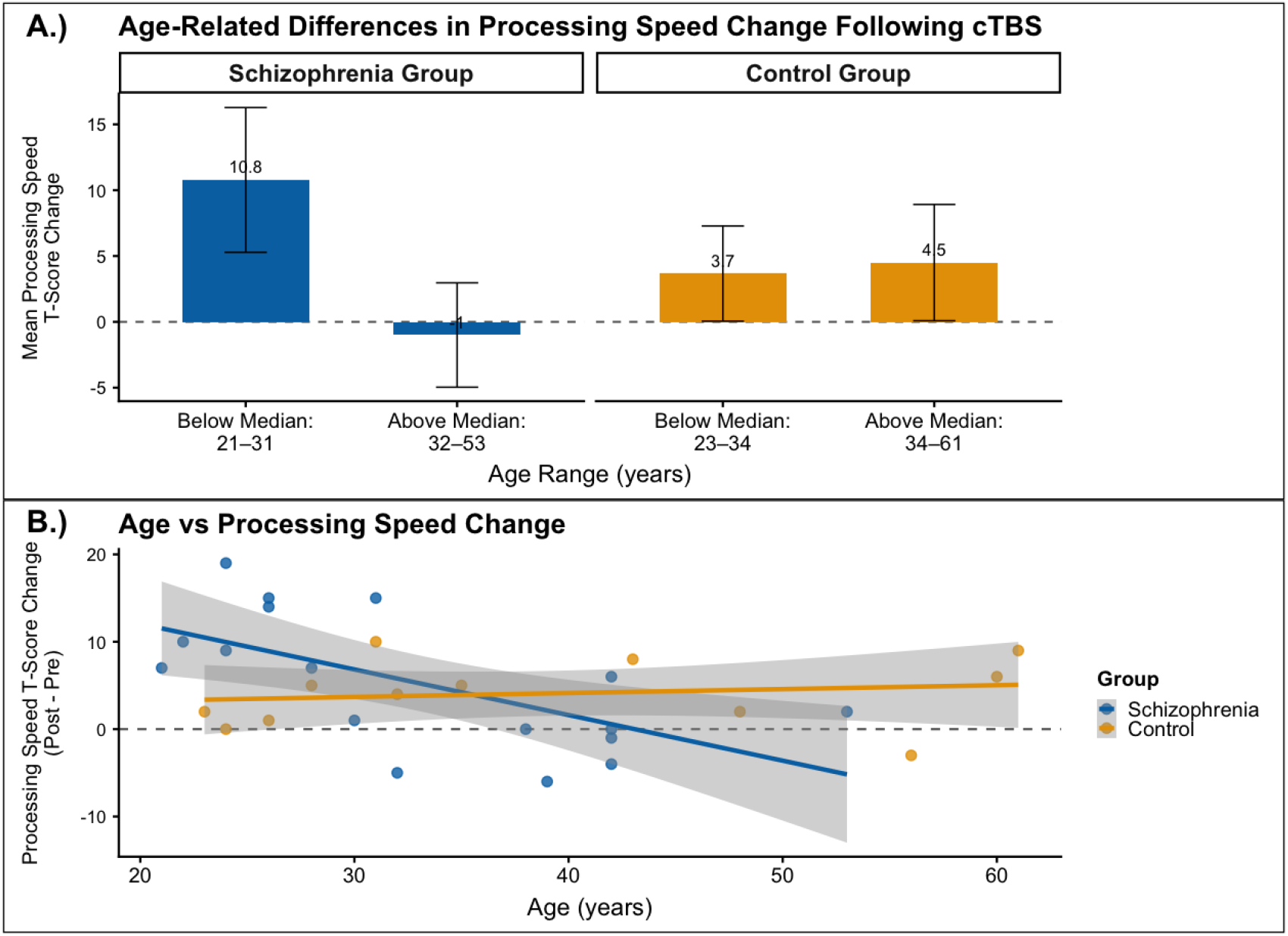
In schizophrenia, there is an age by diagnosis response interaction for processing speed change following cTBS. Twenty-nine participants (17 schizophrenia, 12 non-psychosis controls) received five accelerated sessions of DMN-targeted cTBS with pre-post processing speed assessment. In a linear regression model predicting processing speed change based on age, group, and the age*group interaction (F(3,25)=4.34, p=0.014), group (Estimate=20.15, SE=6.92, t=2.91, p=.0075) and the age*group interaction (Estimate=-0.57, SE=0.19, t—3.03, p=.0056) were significant predictors of processing speed change. Figure 2A displays the effects of group and age on processing speed improvement, we show processing speed improvement based on age below or above the group median (median age 31y schizophrenia, 34y non-psychosis controls). In schizophrenia (blue), age below the median is associated with processing speed improvement, which is not true for participants with age above median. This age effect was not present in the non-psychosis control group (gold), where both age groups show similar improvements in performance.

### DMN Connectivity Change Associated with Processing Speed Change in Schizophrenia

In a linear regression model testing the relationship between processing speed change in relation to age, group, DMN connectivity change, and the group*DMN connectivity change interaction (F(4,21)=2.25, p=0.098), age (Estimate=-0.31, SE=0.13, t=-2.42, p=.025), DMN connectivity change (Estimate=2.22, SE=1.00, t=2.22, p=.037), and the group*DMN connectivity change interaction (Estimate=-3.08, SE=1.23, t=-2.50, p=0.021) were significantly related to processing speed change. In the schizophrenia group, improved processing speed was associated with reduced DMN connectivity, while in the control group, improved processing speed was associated with increased DMN connectivity (Figure 5).

**Figure 5.**
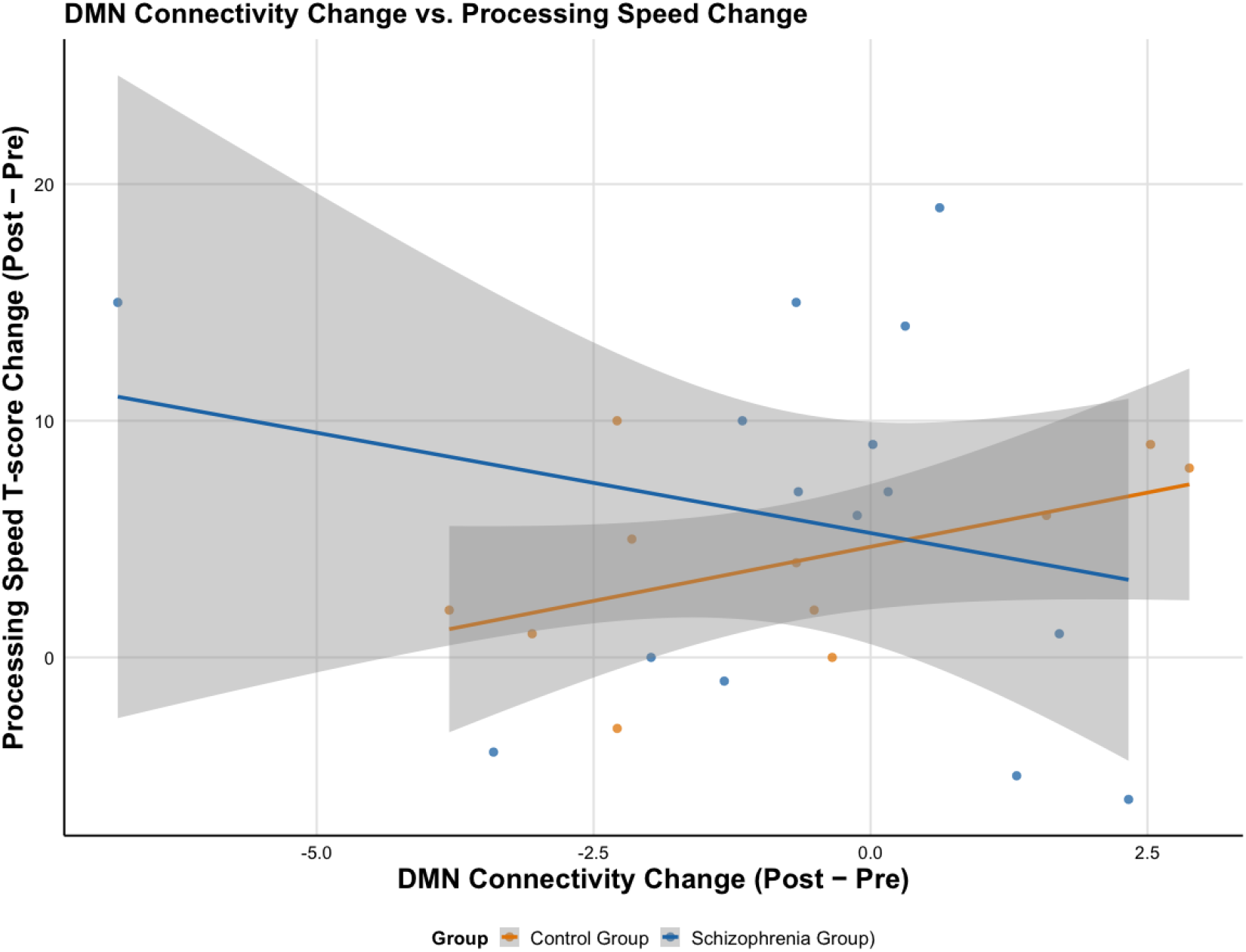
DMN connectivity change is associated with improvements in processing speed in the schizophrenia group and decreased processing speed in the control group. Twenty nine participants (17 schizophrenia, 12 non-psychosis controls) received five accelerated sessions of DMN-targeted cTBS with pre-post resting-state neuroimaging and processing speed assessment. In a linear regression model testing processing speed change based on age, group, DMN connectivity change, and the group*DMN connectivity change interaction (F(4,21)=2.25, p=0.098, age (Estimate=-0.31, SE=0.13, t=-2.42, p=.025), DMN connectivity change (Estimate=2.22, SE=1.00, t=2.22, p=.037), and the group*DMN connectivity change interaction (Estimate=-3.08, SE=1.23, t=-2.50, p=0.021) was associated with processing speed change. In the schizophrenia group, reduced DMN connectivity is associated with improved processing speed. In non-psychosis controls, increased DMN connectivity is associated with improved processing speed, suggesting an opposite mechanistic effect for processing speed improvements between groups.

## Discussion

Although cognitive deficits are consistent, detrimental contributors to poor functional outcomes in schizophrenia, there are no approved pharmacologic treatments for cognitive impairment. In this manuscript, we compared single and multiple sessions of DMN-targeted TMS to establish the efficacy of multiple sessions of TMS in improving processing speed in schizophrenia. We observed that single sessions of DMN-targeted cTBS delivered at 80% AMT did not change processing speed in schizophrenia while accelerated, multi-session DMN-targeted cTBS at 100% AMT significantly improved processing speed. We also observed an age-dependent relationship of processing speed change in schizophrenia, where younger participants experienced greater improvement than older participants. Decreased DMN connectivity correlated with improvement in processing speed in schizophrenia, while increased DMN connectivity corelated with improvement in processing speed in controls. These findings suggest a novel DMN-targeted cTBS treatment for processing speed impairment in schizophrenia.

Single session DMN-targeted TMS delivered at 80% AMT did not change processing speed for iTBS, cTBS, or sham groups. Previous literature has established single session TMS as a helpful method for understanding the short-term mechanistic changes occurring following TMS (23). However, research largely suggests that single sessions of TMS are not strong enough to modulate network-level connectivity needed to produce a behavioral effect (24,25). Therefore, it is not surprising that a single session of TMS did not elicit processing speed change.

During the accelerated, multi session TMS study, cTBS was administered to the DMN at 100% AMT for five sessions. With multiple sessions and increased intensity, we observed a clear improvement in processing speed in the schizophrenia group that we did not see in the single-session study. This supports the notion that five sessions of TMS treatment are adequate to produce a behavioral effect lasting multiple days while single-session TMS treatment was not. Given processing speed was measured with identical MATRICS tasks administered at two points in time, it is important to acknowledge the possibility of a practice effect. However, previous research has established that the BACS processing speed task does not have a significant practice effect (p=0.46, d=0.03) (16), making it an ideal task for multi-visit clinical trials. Furthermore, we do not observe a practice effect during the single-session study where participants received identical cognitive testing approximately one hour apart. In the accelerated study, cognitive testing was administered approximately seven days apart, which would have allowed much more time for memory decay. Therefore, it is unlikely that the improvements observed after multi-session TMS are attributed to a practice effect.

We hypothesized that reducing DMN connectivity through multiple sessions of DMN-targeted cTBS would improve processing speed in schizophrenia. Normal cognitive function requires suppression of DMN activity, a function that is impaired in schizophrenia (7). As expected, our results demonstrated that reduced DMN connectivity was associated with improvement in processing speed in the schizophrenia group. Interestingly, reductions in DMN connectivity in non-psychosis controls predicted *worse* performance on processing speed tasks, suggesting that there may be an ideal level of DMN connectivity to support cognitive performance. Similar trends have been observed with creative thinking in cognition, with an inverted U representing an ideal balance of DMN-ECN connectivity to promote optimal performance on tasks requiring creativity (26). Other research demonstrates that DMN hyperconnectivity leads to cognitive impairments such as slowed information processing while DMN hypoconnectivity leads to impairments such as reduced attentional ability (27,28). Taken together, current literature implies an optimal DMN connectivity range which supports healthy cognitive function.

In the accelerated, multi-session TMS study, we observed an age by diagnosis interaction in which younger participants demonstrated greater improvement in processing speed than older participants in the schizophrenia group. However, age did not correlate with performance in the control group. This finding is in alignment with the accelerated aging hypothesis of schizophrenia which suggests that the cognitive decline experienced by individuals with schizophrenia mirrors the cognitive decline experienced by control populations, but several years earlier (15,29,30). This hypothesis would explain why older adults with schizophrenia did not experience cognitive change while younger adults clearly improved. An alternative explanation may be that younger individuals with schizophrenia have had less exposure to antipsychotic medications, which have been associated with cognitive impairment in psychosis (31,32). Future studies examining biomarkers of aging alongside cognition in schizophrenia are needed to determine if a mechanistic explanation exists beneath this age-related behavioral observation.

This study has several major strengths. First, this study used rigorous study designs applying personalized TMS interventions with neuronavigation and pre-post neuroimaging to demonstrate *both* clinical benefit *and* its underlying mechanism, which is exceptionally rare in the field of psychiatric neuroscience. Application of single TMS sessions in a within-subjects crossover design with dense phenotyping (140 min of resting-state imaging per participant) and use a personalized, network-targeted TMS intervention, particularly in a highly symptomatic population, are innovative. Increasingly, evidence suggests that the heterogeneity in TMS response may be related to individual differences, so the field has moved toward personalized neuromodulation targets. The use of an accelerated TMS protocol in schizophrenia is also novel. Ours is the first study to demonstrate that a personalized, network-targeted TMS intervention improves processing speed in schizophrenia and that this improvement is mediated by changes in DMN connectivity.

Our study also has several important limitations. The accelerated, multi-session cTBS study does not have a sham control group. Therefore, we cannot directly compare the efficacy of our protocol’s active treatment to an inactive treatment. To address this, we compared the effects of single and multiple TMS sessions, which confirmed the superiority of multi-session DMN-targeted cTBS on processing speed. Similarly, the single-session study does not include a control sample, so we are not able to examine processing speed changes after a single session in a non-psychosis population. Furthermore, our sample size was limited to 10 participants in the single session TMS study, and 29 participants in the accelerated, multi-session cTBS study. Future studies are needed to replicate this finding in larger, sham-controlled crossover trials of networked targeted cTBS.

In summary, our findings suggest that accelerated, multi-session cTBS targeted to the DMN improves processing speed in schizophrenia, supporting evidence for a novel treatment for the leading cause of cognitive impairment in this population.

## Supporting information

Supplemental figures and expansion of methods.

## Data Availability

Data available upon reasonable request.

