## Supplemental figures and expansion of methods. for "Accelerated DMN-Targeted cTBS Improves Processing Speed Deficits in Schizophrenia"

**Supplemental Material**

**Methods**

***Single-Session DMN-Targeted TMS in Schizophrenia***

*Participants:* Diagnosis of schizophrenia or schizoaffective disorder was confirmed by DSM-V SCID interview (8) and clinical information obtained from outpatient psychiatric providers. For one month prior to enrollment, individuals received outpatient care, with no hospitalizations or changes to their psychiatric medication regimens. Individuals were excluded if they had DSM-V intellectual disability, substance use disorder (other than nicotine) in the past 3 months, a progressive or genetic neurologic disorder, history of significant head trauma, history of seizures or neurosurgical procedures, implanted devices, gross organic pathology on neuroimaging, contraindications to MRI or TMS, or current pregnancy. All participants provided written informed consent in accordance with the Beth Israel Deaconess Medical Center Institutional Review Board.

*Cognitive Assessments:* Prior to receiving TMS, the Brief Assessment of Cognition in Schizophrenia Digit Symbol Coding (BACS processing speed) task was administered to all participants. The BACS processing speed task is a sub-test of the processing speed domain of the Measurement and Treatment Research to Improve Cognition in Schizophrenia (MATRICS). The MATRICS is a validated, standard cognitive battery developed to detect cognitive deficits in Schizophrenia (16). The BACS processing speed task requires participants to match numbers with an associated nonsense symbol using paper and pen. Participants are scored on the number of correct matches completed during a 90 second period. The MATRICS online scoring program transforms raw scores into a corrected t-score to account for age and gender (17). After TMS administration (i.e., approximately two hours after the first test), participants performed the BACS Digit Symbol Coding task again.

*MRI Acquisition and Data Processing:* Imaging data were collected on a Siemens 3.0-T MRI system (Munich, Germany). Participants each completed 7 MRI scans, including at baseline and immediately before and after each TMS session (Figure 1B). A total of 270 min of imaging data was collected per participant, including 140 min of resting-state imaging. Briefly, 1-mm$^3$ T1-weighted anatomical scans and multiple 10-minute functional runs were acquired (TR 650ms, TE

34.80ms, flip angle 50 degrees, multiband acceleration factor 8, field of view = 207mm, 64 slices, 3-mm$^3$ voxels, anterior to posterior phase-encoded). Anatomical images were segmented into GM, WM, and CSF with the Computational Anatomy Toolbox 12 (CAT12, version 12.5; http://www.neuro.uni-jena.de/cat/). Resting-state scans were preprocessed in SPM12 (https://github.com/vuIIS/vuiis-cci-info?tab=readme-ov-file#citing-xnatdax) and (1) realigned to a mean scan, (2) coregistered with the native space structural scan, then (3) underwent resting-state denoising procedures: bandpass filter (0.01–0.1 Hz), regression of CSF, WM, and mean GM signal, regression of 12 motion parameters (6 translation and rotation parameters and their first derivative). All resting-state scans went through a quality assurance procedure that included calculating framewise displacement (FD) and temporal signal to noise ratio (tSNR). Scans with mean FD>0.5 or tSNR<5th percentile of the sample distribution were excluded. After quality control, there were pre- and post-TMS scans for 8 cTBS, 9 iTBS, and 6 sham sessions across 9 subjects.

*TMS Protocol:* Individuals participated in a randomized, controlled, crossover study of three single sessions of theta-burst stimulation (Figure 1B and Supplemental Figure 1) applied to an individualized DMN target (see *Individualized DMN Target* below and Supplemental Figure 3) with neuroimaging collected immediately before and after each TMS session. Baseline anatomical and functional MRIs were used in a Brainsight frameless stereotaxic system (Rogue Research, Montreal, Canada) to target an individualized left lateral parietal DMN region. Frameless stereotaxy was used during all stimulation sessions to monitor the position of the coil throughout TMS administration. Participants were seated in a chair with their head tilted downward. Individuals received three single sessions of TMS on three separate days, separated by at least 2 days (48 hours) to avoid any carryover effect (median 6.5 days, mean 13.8 days (SD 22.2), range 2-96 days). Active motor threshold (AMT) was determined prior to the first TMS session. Participants received one session of iTBS (600 pulses, 100% AMT), one session of cTBS (600 pulses, 80% AMT), and one session of sham (coil flipped 180 degrees using 100% AMT iTBS protocol, 600 pulses). TMS session order was randomly assigned. TMS was applied using a MagPro X100 stimulator and an active figure-of-8 coil (Cool B65, MagVenture, Denmark) held tangentially to the scalp with the handle at 45 degrees. TMS was applied in the standard theta-burst pattern described by Huang et al. (3 pulses at 50-Hz repeated at a rate of 5-Hz) (9).

Participants were blinded to TMS stimulation type. To assess the integrity of the blind, participants were asked after each TMS session what type of TMS they thought they received.

*TMS Protocol Motor Threshold Determination:* Participants had motor threshold determination at their first TMS visit. Single pulse and repetitive stimulation was performed with a MagPro stimulator (MagVenture) equipped with a biphasic figure-of-eight coil. To obtain an active motor threshold, single pulses were used in the following manner: Electromyographic activity (EMG) was recorded using surface electrodes attached to the skin to measure motor evoked potentials (MEP) during the motor threshold assessment. The TMS coil was placed on the scalp. Single TMS pulses were applied over the hand area of the left motor cortex and individually localized for each participant based on the optimal position for eliciting a motor evoked potential. Neuronavigation (Brainsight, Rogue Research, Inc.) was used to record the motor 'hot-spot.' Resting and active motor threshold (RMT; AMT) were obtained by following recommendations from the International Federation of Clinical Neurophysiology.

*TMS Neuroimaging Analysis - Calculation of TMS Target DMN Connectivity Values:* For the Single-Session TMS and Accelerated, Multi-Session cTBS analyses, we calculated average connectivity to the DMN from the TMS target (left lateral parietal DMN) by extracting time courses from eight standard DMN nodes (posterior cingulate/precuneus, medial prefrontal, right lateral parietal, left inferior temporal, right inferior temporal, medial dorsal thalamus, right posterior cerebellum, left posterior cerebellum) and averaging the connectivity values between the TMS target and the eight other DMN nodes for each participant.[6] We calculated individual values of DMN functional connectivity by placing 6mm spheres at coordinates corresponding to standard DMN nodes ([15] for coordinates). The BOLD signal time courses from the TMS target (left lateral parietal DMN) and the other DMN regions of interest (ROIs) were correlated with each other and z-transformed to generate ROI-to-ROI connectivity values. The mean DMN connectivity from the TMS target was generated for each participant by averaging connectivity values of these eight network edges.

***Accelerated Multi-Session DMN-Targeted TMS in Schizophrenia***

*Participants:* Diagnosis of schizophrenia or schizoaffective disorder was confirmed by DSM-V SCID interview (8) and clinical information obtained from outpatient psychiatric providers. For one month prior to enrollment, individuals received outpatient care, with no hospitalizations or changes to their psychiatric medication regimens. Individuals were excluded if they had DSM-V intellectual disability, substance use disorder (other than nicotine) in the past 3 months, a progressive or genetic neurologic disorder, history of significant head trauma, history of seizures or neurosurgical procedures, implanted devices, gross organic pathology on neuroimaging, contraindications to MRI or TMS, or current pregnancy. All participants provided written informed consent in accordance with the Vanderbilt University Medical Center Institutional Review Board.

*Cognitive Assessment*: Participants were administered the three tasks comprising the MATRICS Processing Speed Domain including BACS Digit Symbol Coding (BACS processing speed), Category Fluency: Animal Naming, and Trail Making Test: Part A. The BACS processing speed task requires participants to match numbers with an associated nonsense symbol using paper and pen during a 90 second period. The Animal Naming task required participants to orally name as many animals as possible in 60 seconds (20). The Trail Making Test requires participants to draw a continuous line in sequential order for 25 randomly placed dots on a sheet of paper (21). The MATRICS online scoring program provided T-scores for each sub-test as well as a composite Processing Speed T-score derived from a participant's overall performance on the three tasks. Participants completed the MATRICS tasks during the pre-TMS and post-TMS MRI visits, with at least four days in between tests (median 7 days, mean 7.79 days (SD 2.57), range 4-14 days).

*TMS Protocol:* Individuals received single sessions of theta-burst stimulation applied to an individualized DMN target with neuroimaging collected immediately before and after each session. Individuals received one session of iTBS (600 pulses, 100% Active Motor Threshold, AMT), cTBS (600 pulses, 80% AMT as per (18)), and sham (coil flipped 180 degrees using 100% AMT iTBS protocol, 600 pulses) on three separate days, separated by at least 2 days to avoid carryover effect (median 6.5 days, mean 13.8 days (SD 22.2), range 2-96 days). TMS was applied using a MagPro X100 stimulator and an active figure-of-8 coil (Cool B65, MagVenture,

Denmark) held tangentially to the scalp with the handle at 45 degrees. TMS was applied in the standard theta-burst pattern (3 pulses at 50-Hz repeated at a rate of 5-Hz) (19). Order was randomized, and participants were blinded to stimulation type.

*MRI Acquisition and Data Processing:* Imaging data were collected on 3.0-T Philips Intera Achieva MRI scanner (Philips Healthcare, Andover, MA). Participants completed MRI scans before and after the TMS intervention. As the durability of 5 TMS sessions is unknown, post-TMS scans were scheduled 3 to 7 days after TMS per protocol. Briefly, 1-mm$^3$ T1-weighted anatomical scans and multiple 10-minute functional runs were acquired (TR 2000ms, TE 28.0ms, flip angle 90 degrees, field of view = 240mm, 38 slices, 3-mm$^3$ voxels, anterior to posterior phase-encoded). Anatomical images were segmented into GM, WM, and CSF with the Computational Anatomy Toolbox 12 (CAT12, version 12.5; http://www.neuro.uni-jena.de/cat/). Resting-state scans were preprocessed in SPM12 (https://github.com/vuIIS/vuiis-cci-info?tab=readme-ov-file#citing-xnatdax) and (1) realigned to a mean scan, (2) coregistered with the native space structural scan, then (3) underwent resting-state denoising procedures: bandpass filter (0.01–0.1 Hz), regression of CSF, WM, and mean GM signal, regression of 12 motion parameters (6 translation and rotation parameters and their first derivative). All resting-state scans went through a quality assurance procedure that included calculating FD and tSNR. Scans with mean FD > 0.6 or tSNR lower than the 5th percentile of the sample distribution were excluded from analysis. After quality control, there were pre-/post-TMS scans for 15 schizophrenia participants and 11 non-psychosis control participants.

*TMS Protocol Motor Threshold Determination:* Participants had motor threshold determination at their first TMS visit. Single pulse and repetitive stimulation was performed with a MagPro stimulator (MagVenture) equipped with a biphasic figure-of-eight coil. To obtain an active motor threshold, single pulses were used in the following manner: Electromyographic activity (EMG) was recorded using surface electrodes attached to the skin to measure motor evoked potentials (MEP) during the motor threshold assessment. The TMS coil was placed on the scalp. Single TMS pulses were applied over the hand area of the left motor cortex and individually localized for each participant based on the optimal position for eliciting a motor evoked potential. Neuronavigation (Brainsight, Rogue Research, Inc.) was used to record the motor 'hot-spot.' Resting and active motor threshold (RMT; AMT) were obtained by following recommendations from the International Federation of Clinical Neurophysiology.

*TMS Neuroimaging Analysis - Calculation of TMS Target DMN Connectivity Values:* For the Single-Session TMS and Accelerated, Multi-Session cTBS analyses, we calculated average connectivity to the DMN from the TMS target (left lateral parietal DMN) by extracting time courses from eight standard DMN nodes (posterior cingulate/precuneus, medial prefrontal, right lateral parietal, left inferior temporal, right inferior temporal, medial dorsal thalamus, right posterior cerebellum, left posterior cerebellum) and averaging the connectivity values between the TMS target and the eight other DMN nodes for each participant.[6] We calculated individual values of DMN functional connectivity by placing 6mm spheres at coordinates corresponding to standard DMN nodes ([15] for coordinates). The BOLD signal time courses from the TMS target (left lateral parietal DMN) and the other DMN regions of interest (ROIs) were correlated with each other and z-transformed to generate ROI-to-ROI connectivity values. The mean DMN connectivity from the TMS target was generated for each participant by averaging connectivity values of these eight network edges: Posterior Cingulate Cortex, Medial Prefrontal Cortex, Right Lateral Parietal, Left Inferior Temporal, Right Inferior Temporal, Medial Dorsal Thalamus, Left Posterior Cerebellum, and Right Posterior Cerebellum.

***Individualized DMN Targeting****:* The DMN target was identified using the same methods for the Single-Session TMS and Accelerated Multi-Session TMS studies. For our DMN target, we selected the left lateral parietal DMN, as it is a DMN region that is readily identifiable across all individuals and has been successfully used to modulate DMN connectivity with TMS (11). To identify an individualized DMN map for TMS targeting, a standard DMN template (12) was warped into native space and applied to the participant's baseline or pre-TMS scan. In each participant, the resultant connectivity maps yielded a correlation cluster in the left posterior inferior parietal lobule (IPL, Supplemental Figure 3). A target was then placed in the averaged center of the left posterior IPL correlation cluster (formed from the overlay of the left posterior IPL clusters derived from the connectivity maps) on the cortical surface using Brainsight neuronavigation software (Rogue Research, Inc.) An individualized TMS target was selected in the left parietal region of the DMN and used as the TMS target for all TMS sessions.

### Results

#### *Single-Session DMN-Targeted TMS*

*Single-Session DMN-Targeted TMS is Safe and Well-Tolerated in Schizophrenia*

TMS was safe and well-tolerated. All 10 participants who were randomized completed all three TMS/fMRI visits. No serious adverse events were observed. Participants endorsed the following side effects: headache or neck pain (n=4) and trouble concentrating (n=5). One participant experienced hypomania that responded to medication adjustment. Four participants reported lower anxiety, and one participant reported improved mood.

After each TMS single session, participants were asked what type of TMS they thought they received: cTBS, iTBS, or sham. Of 30 total TMS sessions, in 14 of them (46.67%) the participant correctly guessed the type of TMS they received. Of 10 sham sessions, in 4 of them (40%), participants correctly guessed that they were receiving sham.

#### *Accelerated Multi-Session DMN-Targeted TMS*

*Accelerated Multi-Session DMN-Targeted TMS is Safe and Well-Tolerated in Schizophrenia*

TMS was safe and well-tolerated. All 12 participants received all 5 sessions of cTBS and underwent pre-/post-TMS neuroimaging. No serious adverse events were observed. One participant reported mild anxiety and low mood after the first TMS session and slight irritability the day after TMS.

### Supplemental Tables & Figures:

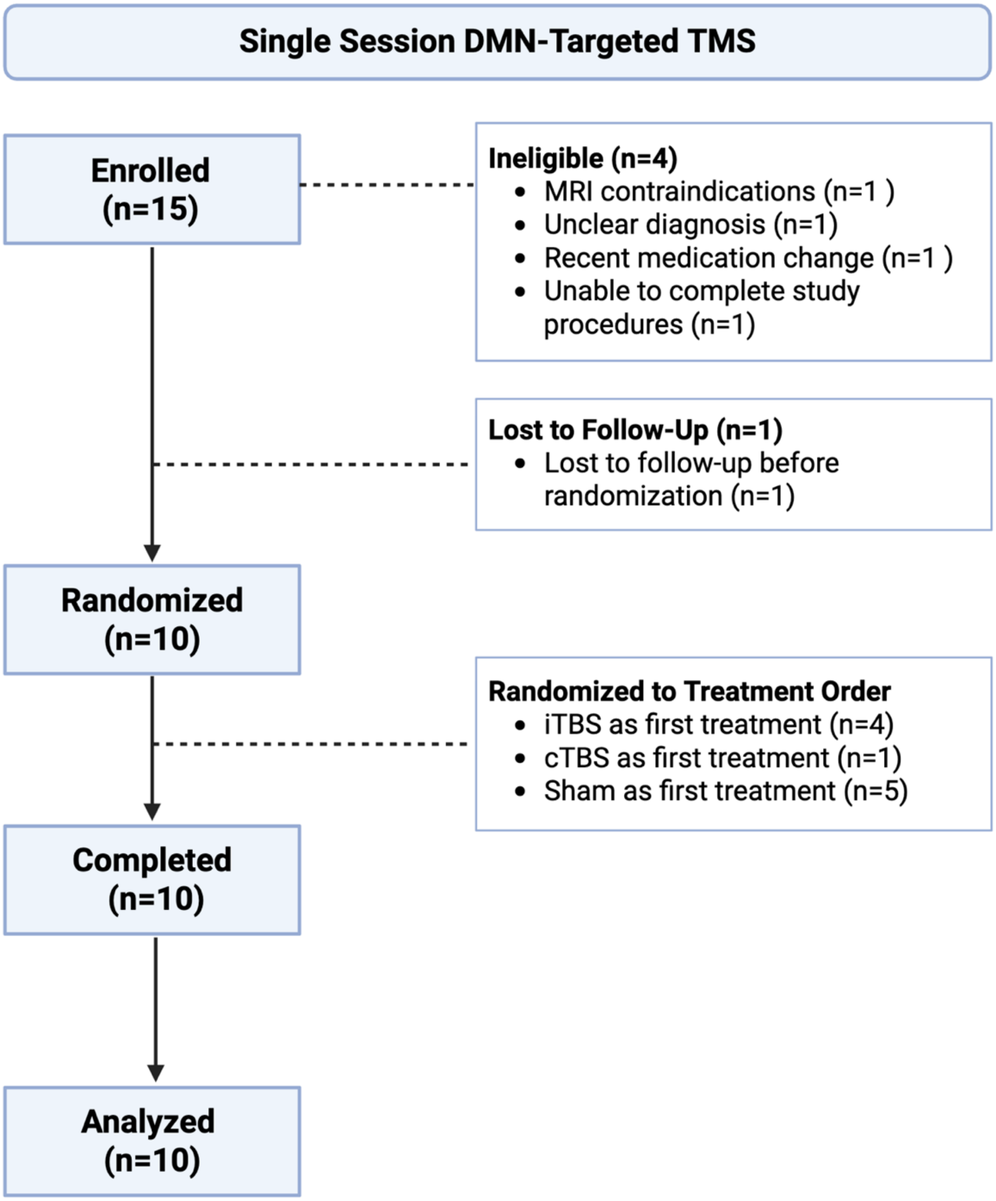


**Supplemental Figure 1. Single-Session DMN-Targeted TMS CONSORT Diagram.** Fifteen participants with schizophrenia or schizoaffective disorder aged 18-65 were enrolled in this randomized, sham-controlled crossover study. Ten participants completed the study and provided data for analysis.

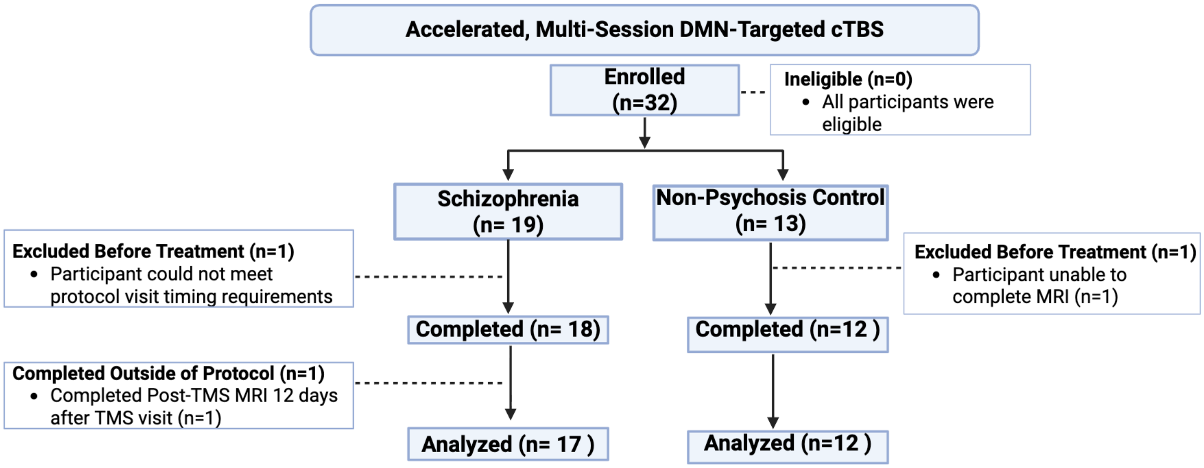


**Supplemental Figure 2. Accelerated, Multi-Session DMN-Targeted cTBS CONSORT Diagram.** Nineteen participants with schizophrenia or schizoaffective disorder aged 18-65 were enrolled in this open-label study. Eighteen participants completed the study procedures, and one participant was excluded from analysis due to completion outside of protocol requirements. Thirteen non-psychosis control participants were enrolled in this open-label study. One participant was unable to complete an MRI and was therefore excluded from the study prior to cTBS treatment. All non-psychosis control participants were included in analysis.

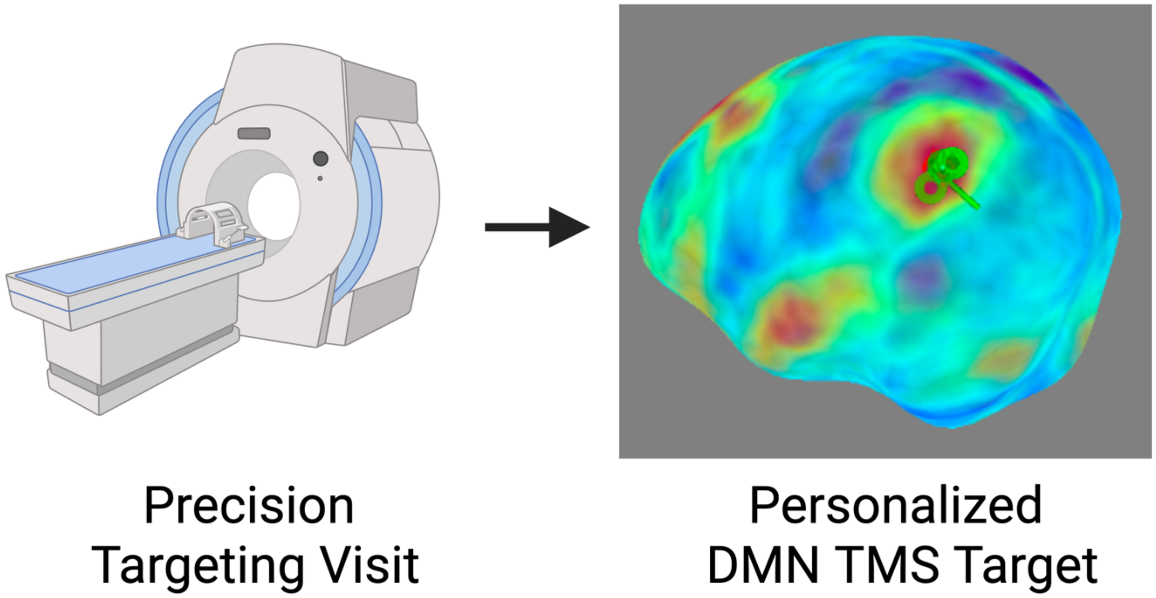


**Supplemental Figure 3. Individualized Default Mode Network TMS Target***:* The Default Mode Network (DMN) target was identified using the same methods for the Single-Session TMS and Accelerated Multi-Session TMS studies. For our DMN target, we selected the left lateral parietal DMN, as it is a DMN region that is readily identifiable across all individuals and has been successfully used to modulate DMN connectivity with TMS (11). To identify an individualized DMN map for TMS targeting, a standard DMN template (12) was warped into native space and applied to the participant's baseline or pre-TMS scan. The participant scan and DMN mask were then warped back into standard MNI space. In each participant, the resultant connectivity maps yielded a correlation cluster in the left posterior inferior parietal lobule. A target was then placed in the averaged center of the left posterior inferior parietal lobule correlation cluster (formed from the overlay of the left posterior inferior parietal lobule clusters derived from the connectivity maps) on the cortical surface using Brainsight neuronavigation software (Rogue Research, Inc.) An individualized TMS target was selected in the left parietal region of the DMN and used as the TMS target for all TMS sessions. Created with BioRender.com.

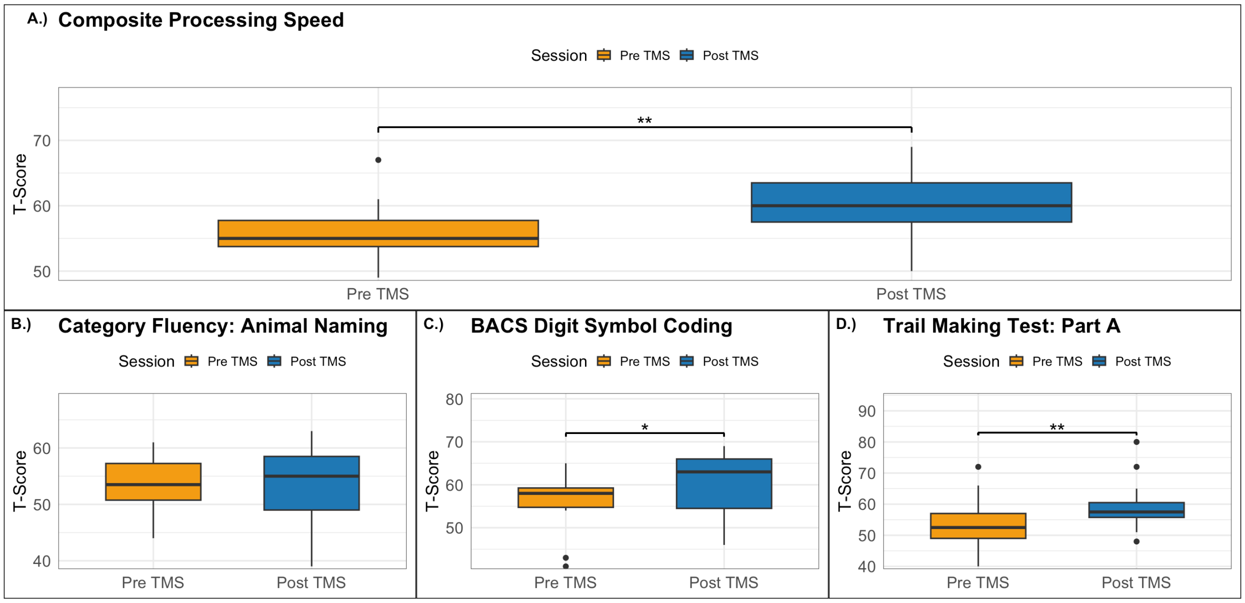


**Supplemental Figure 4. Accelerated, Multi Session cTBS improves overall processing speed in a non-psychosis control sample.**

Twelve individuals without a psychotic disorder received five accelerated sessions of DMN-targeted cTBS with pre-post assessment of processing speed. Processing speed was assessed using three individual tests: 1) Category Fluency: Animal Naming; 2) BACS Digit Symbol Coding and 3) Trail Making Test (TMT): Part A, which were compiled to create a composite processing speed t-score corrected for age and gender. Composite processing speed score (p= 0.0038, Supplemental Figure 4A), Digit Symbol Coding score (p=0.0207), Supplemental Figure 4C), and TMT (p= 0.002, Supplemental Figure 4D) significantly improved in the schizophrenia group. Animal Fluency (p=0.5240195, Supplemental Figure 4B) revealed a non-significant numerical increase in median.

**Supplemental Table 1. Demographics**

| | **Single Session TMS** | | **Accelerated Multi-Session cTBS** | |
|---|---|---|---|---|
| | **Psychosis** (n=11) | **Psychosis** (n=19) | *p* | **Control** (n=13) |
| **Age, y (SD)** | 38.73 (13.70) | 32.1 (9.1) | 0.0634 | 40 (14.1) |
| **Sex, n (% Female)** | 8M/3F (27.3) | 16M/3F (15.8%) | 0.219 | 5F/8M (38.5%) |
| Black/African American | 2/11 (18.2) | 9 (47.4%) | 0.0237 | 1 (7.7%) |
| Native American | 1/11 (9.1) | 0 (0%) | 0.406 | 1 (7.7%) |
| White | 8/11 (72.3) | 8 (42.1%) | 0.166 | 9 (69.2%) |
| Native Hawaiian or Other Pacific Islander | 0/11 (0.0) | 0 (0%) | - | 0 (0%) |
| Asian | 0/11 (0.0) | 0 (0%) | - | 0 (0%) |
| Multiracial | 0/11 (0.0) | 1.00 | 1.00 | 1 (7.7%) |
| Other, not specified | 0/11 (0.0) | 0 (0%) | 0.406 | 1 (7.7%) |
| Unknown/missing | 0/11 (0.0) | 0 (0%) | - | 0 (0%) |
| PANSS Total (SD) | 55.27 (11.84) | - | - | - |
| BPRS Total (SD) | - | 37.39 (6.37) | 0.009 | 31.62 (6.46) |
| AMT | 40.9 (6.82) | 48.33 (19.37) | 0.292 | 41.91 (8.68) |
| cTBS Intensity | 80% AMT | - | - | 100% AMT |
| iTBS Intensity | 100% AMT | - | - | - |

**Note**: In the single-session DMN-targeted TMS study, 15 participants were enrolled, 4 were ineligible, and 11 completed baseline assessments. In the accelerated, multi-session DMN-targeted cTBS study, 32 participants were enrolled, all of whom were eligible. Two participants withdrew (1 schizophrenia, 1 control) after completing baseline assessments. One participant was excluded from all analyses due to protocol nonadherence. Data for analysis included 17 schizophrenia participants and 12 controls.
